# Associations of plasma omega-3 fatty acid levels and reported fish oil supplement use with depression and anxiety: A cross-sectional analysis from the UK Biobank

**DOI:** 10.1101/2025.04.04.25325253

**Authors:** William S. Harris, Jason Westra, Nathan L. Tintle, Prasad P. Devarshi, Ryan W. Grant, Susan Hazels Mitmesser

## Abstract

The role that marine omega-3 polyunsaturated fatty acids (PUFAs) may play in reducing the risk for developing depression and/or anxiety is unclear. The present study examined the relationships between plasma levels of total omega-3 PUFAs, docosahexaenoic acid (DHA) and the non-DHA omega-3 PUFAs with medical-record documented depression and/or anxiety (both historical and recent, within the last 12 months) in the UK Biobank. The associations of these outcomes with the reported use of fish oil supplements (FOS) were also examined. Data from 258,354 participants who had data on plasma omega-3 PUFA levels and all covariates were used for the biomarker-based analyses, and data from 468,145 people who reported FOS use at baseline were used in the latter analysis. We found that all three omega-3 PUFA metrics were inversely associated with a history of both depression and anxiety. Specifically, risk for the former outcome was between 15% and 33% lower in Q5 vs Q1, and for the latter outcome, between 19% to 22% lower comparing Q5 with Q1. Risk for recent depression was 29% and 32% lower (Q5 vs Q1) for total omega-3 PUFAs and for non-DHA, respectively. FOS use was associated with a 9-10% lower risk for a history of depression and anxiety, respectively, and a 20% lower risk for recent anxiety. In conclusion, we found evidence that higher levels of omega-3 PUFAs may play a protective role in depression and anxiety.

## INTRODUCTION

Depression is a mental health condition that involves a prolonged period of sadness, loss of interest, and difficulty with daily life. Its estimated prevalence in 2020 in the US was about 18%[1]. Some cross-sectional and prospective cohort studies have suggested that a low level of omega-3 polyunsaturated fatty acids (PUFAs), especially DHA, in the blood/tissues is associated with increased risk for depression[2]. Furthermore, a meta-analysis of randomized clinical trials (RCTs) also supports this conclusion[3]. However, others have suggested no relationship. For example, a sub-study of VITAL (an RCT testing the effects of 840 mg/d EPA+DHA ethyl esters for about 5 years in >25,000 adults free of cardiovascular disease and cancer at baseline,) entitled VITAL-Depression Endpoint Progression[4] examined the effects of omega-3 PUFA on incident depression in >18,000 participants free of depression at baseline. Compared with placebo, treatment with omega-3 supplements yielded mixed results, with a small but statistically significant increase in risk of depression or clinically relevant depressive symptoms but no difference in mood scores. The authors concluded that “These findings do not support the use of omega-3 supplements in adults to prevent depression.” On the other hand, in another study treatment of individuals with depression with 3.2 g of omega-3 PUFAs for 12 weeks had favorable results, with the authors concluding, “These findings suggested that monotherapy of omega-3 PUFAs could improve depression and potentially serve as an alternative option for patients with major depressive disorder (MDD).”[5] Thus, the relationships between omega-3 and depression remains unclear.

The situation is similar for anxiety, which is defined as an emotion characterized by feelings of fear, dread, tension, and worried thoughts. Like depression, anxiety levels continue to increase in the U.S. and globally, with a marked rise during and after the global pandemic. Both biomarker-based case control studies[6] as well as meta-analyses of RCTs[7] support a beneficial role of omega-3 PUFAs in anxiety, but there are notable exceptions[8]. Hence, like depression, the role that omega-3 PUFAs may play in either the prevention or treatment of anxiety disorders requires further examination.

To attempt to clarify these relationships, we examined the cross-sectional relationships between plasma levels of omega-3 PUFAs and DHA specifically (as well as reported fish oil use) and depression and anxiety in the UK Biobank.

## METHODS

### Sample

The UKBB is a prospective, population-based cohort of 502,411 individuals, 40-70y, recruited in the UK between April 2007 and December 2010 (Sudlow et al. 2015). UKBB has ethical approval (Ref. 11/NW/0382) from the Northwest Multi-centre Research Ethics Committee as a Research Tissue Bank, and the University of South Dakota Institutional Review Board approved the use of these de-identified, publicly available data for research purposes (IRB-21-147). Within the cohort, a random set of 258,354 participants had available FA data after exclusion of those missing covariates, and those who withdrew from the UK Biobank project. In some analyses, we consider a sample of 468,145 people with answers to a question on fish oil use and complete covariate data.

### Exposures

The primary exposures in this study were plasma DHA (DHA%), total n3 PUFA (%) and non-DHA n3 PUFA (%), the latter defined as total n3 PUFA minus DHA, as measured by NMR (Nightingale Health Plc, Helsinki, Finland). An estimated Omega-3 Index (erythrocyte EPA+DHA; eO3I) was derived from an inter-laboratory experiment as described by Schuchardt et al.[9] Each was expressed as a percent of total FAs[10]. Self-reported regular fish oil supplement (FOS) use was collected by touch screen answers at baseline.

### Outcomes

Lifetime prevalence of depression or anxiety was assessed by any ICD10 code related to these diagnoses in the subjects’ medical records at the time of fatty acid measurement. Recent prevalence of depression or anxiety was assessed by any ICD10 coded depression or anxiety diagnosis within 12 months of the fatty acid measurement).

### Covariates

We adjusted for several demographic, behavioral, biomarker and medical history variables: age, biological sex (Male/Female), self-reported race/ethnicity (Asian, Black, White, other), marital status (married/not), Body Mass Index (BMI), smoking (pack years), self-reported alcohol consumption (Rarely, Monthly, 1-2x/week, 3-4x/week, Daily), Townsend Deprivation Index, education status (college, associates, SE, None), self-reported exercise (quartiles of moderate-to-vigorous exercise minutes/week), self-reported multivitamin use (yes/no), linoleic acid (LA) levels and non-LA omega-6 levels. Additional details on covariates along with corresponding UKBB variable IDs are provided in **Supplemental Table 1**.

### Statistical Analysis

Sample characteristics are summarized using standard statistical methods (e.g., means, SDs, %s). Logistic regression models were fit when predicting history of depression, history of anxiety, current depression, and current anxiety (yes/no) with separate models for each of three n3 PUFA exposures and each of the four outcomes in both partially adjusted (age, sex, race/ethnicity; Model 1) and fully adjusted (all variables shown in **Table 1**; Model 2) models. Each exposure was analyzed for its relationship for mental health diagnosis continuously (per standard deviation, SD) and per quintile (Q). Cubic splines were fit using the splines package in R with knots specified at each quintile and were adjusted for all Model 2 covariates. Tests of non-linearity were conducted by comparing spline models vs. models with standard linear terms. For the associations of outcomes with FOS use, the same model covariates were used except for the plasma FA values. Statistical significance was set to 0.05 for all analyses and 95% confidence intervals (CI) are provided where appropriate. R (www.r-project.org) was used for all analyses.

**Table 1.** Demographics of the sample (N=258,354)

| Characteristic | Mean (SD) or % (n) |
| --- | --- |
| Sex – Female | 53.1% (137,237) |
| Age – years | 56.7 (8.1) |
| Ethnicity |  |
| White | 95.5% (246,789) |
| Asian | 1.8% (4,760) |
| Black | 1.3% (3,441) |
| Other | 1.3% (3,364) |
| Marital status – married/partnered | 72.8% (188,069) |
| Employment status |  |
| Employed | 59.1% (152,703) |
| Disabled | 3.4% (8,685) |
| Retired | 34.5% (89,076) |
| Unemployed | 1.6% (4,115) |
| Other | 1.5% (3,775) |
| Educational status |  |
| College | 60.4% (156,123) |
| High school | 22.1% (57,133) |
| Less than high school | 17.5% (45,098) |
| Alcohol consumption<sup>1</sup> |  |
| Daily | 20.4% (52,801) |
| 3-4x/week | 23.5% (60,692) |
| 1-2x/week | 26.2% (67,775) |
| Rarely | 29.8% (77,086) |
| BMI – mg/kg<sup>2</sup> | 27.5 (4.8) |
| Smoking – pack years | 10.6 (15.5) |
| Townsend Deprivation Index | -1.4 (3.0) |
| Exercise -METS<sup>1</sup> |  |
| Lowest quartile | 22.5% (58,070) |
| Second quartile | 23.0% (59,432) |
| Third quartile | 23.1% (59,627) |
| Top quartile | 23.2% (60,047) |
| Linoleic Acid (%) | 28.9 (3.4) |
| Non Linoleic Omega-6 Fatty acids (%) | 9.0 (1.9) |
| DHA (%) | 2.0 (0.7) |
| Non-DHA Omega 3 fatty acids (%) | 2.4 (1.0) |
| Total omega-3 fatty acids (%) | 4.4 (1.6) |
| Self-reported Fish oil consumption | 31.6% (80,912) |

1. Missing data of ~8.2% for this variable.

## RESULTS

The sample size for this cross-sectional study was 258,354 individuals with data on plasma fatty acid levels, the covariates used in the model building and the outcomes of interest. In the aggregate, they were mostly White individuals, over half were women, their mean age was 56 years, and the vast majority were either still working or retired. Over half had post-high school education. About 32% reported that they regularly used FOS (Table 1).

### Associations between plasma omega-3 levels and outcomes

For a lifetime history of depression and anxiety, we found in model 2 that DHA, Total Omega-3, Non-DHA Omega-3 and FOS use were all significantly and inversely associated with history of depression, both linearly (per IQ_5_R) and comparing Q5 vs. Q1. Briefly, relative risk was between 7% and 13% lower for these metrics in linear analyses, and between 15% and 33% lower in Q5 vs Q1 (**Table 2**). For the eO3I, relative risk was 18% lower in each analysis (**Table 3**). For anxiety (which was less prevalent than depression, about 0.5% vs 1%), in linear analyses, total omega-3 levels were associated with a 5% lower risk per IQ_5_R. For total omega-3 and non-DHA, risk for a history of anxiety was 19% to 22% lower comparing Q5 with Q1. For the eO3I, relative risk was 19% lower in Q5 vs Q1.

**Table 2.** Association of omega-3 fatty acid levels with lifetime prevalence of depression and anxiety^1^.

|  |  | History of Depression |  |  |
| --- | --- | --- | --- | --- |
|  |  | Lifetime prevalence | Model 1<sup>2</sup> | Model 2<sup>3</sup> |
| DHA | IQ<sub>5</sub>R | 1.04% | **0.64 (0.60,0.69)\*\*\*** | **0.91 (0.84,0.97)\*\*** |
|  | Q1 | 1.53% | 1.00 | 1.00 |
|  | Q2 | 1.18% | **0.74 (0.66, 0.83)\*\*\*** | 1.00 (0.89,1.12) |
|  | Q3 | 0.97% | **0.60 (0.53, 0.67)\*\*\*** | 0.92 (0.81,1.04) |
|  | Q4 | 0.77% | **0.47 (0.41, 0.53)\*\*\*** | **0.82 (0.71, 0.94)\*\*** |
|  | Q5 | 0.77% | **0.46 (0.41, 0.53)\*\*\*** | **0.85 (0.73, 0.98)\*** |
| Total omega 3 | IQ<sub>5</sub>R | 1.04% | **0.88 (0.86, 0.91) \*\*\*** | **0.93 (0.90, 0.96) \*\*\*** |
|  | Q1 | 1.53% | 1.00 | 1.00 |
|  | Q2 | 1.18% | **0.83 (0.74, 0.93)\*\*** | 0.94 (0.83,1.05) |
|  | Q3 | 0.97% | **0.77 (0.68, 0.86) \*\*\*** | **0.88 (0.78, 0.99)\*** |
|  | Q4 | 0.77% | **0.68 (0.60, 0.76) \*\*\*** | **0.83 (0.73, 0.94)\*\*** |
|  | Q5 | 0.77% | **0.59 (0.52, 0.67) \*\*\*** | **0.74 (0.64, 0.85)\*\*\*** |
| Non DHA Omega 3 | IQ<sub>5</sub>R | 1.04% | **0.92 (0.88, 0.95)\*\*\*** | **0.87 (0.83, 0.92)\*\*\*** |
|  | Q1 | 1.53% | 1.00 | 1.00 |
|  | Q2 | 1.18% | 0.91 (0.81, 1.03) | 0.90 (0.79,1.01) |
|  | Q3 | 0.97% | **0.85 (0.75, 0.96)\*\*** | **0.79 (0.69, 0.89)\*\*\*** |
|  | Q4 | 0.77% | **0.83 (0.73, 0.94)\*\*** | **0.73 (0.64, 0.84)\*\*\*** |
|  | Q5 | 0.77% | **0.77 (0.68, 0.87)\*\*\*** | **0.67 (0.58, 0.77)\*\*\*** |
|  |  | History of Anxiety |  |  |
| DHA | IQ<sub>5</sub>R | 0.50% | **0.76 (0.69,0.83)\*\*\*** | 0.92 (0.83,1.01) |
|  | Q1 | 0.66% | 1.00 | 1.00 |
|  | Q2 | 0.52% | **0.76 (0.64,0.89) \*\*\*** | 0.92 (0.78,1.1) |
|  | Q3 | 0.50% | **0.72 (0.61,0.85) \*\*\*** | 0.96 (0.81,1.15) |
|  | Q4 | 0.40% | **0.57 (0.47,0.68) \*\*\*** | **0.82 (0.67,1.00)\*** |
|  | Q5 | 0.43% | **0.60 (0.51,0.72) \*\*\*** | 0.87 (0.71,1.07) |
| Total omega 3 | IQ<sub>5</sub>R | 0.50% | **0.93 (0.9,0.97) \*\*\*** | **0.95 (0.92,0.99)\*** |
|  | Q1 | 0.61% | 1.00 | 1.00 |
|  | Q2 | 0.48% | 0.78 (0.66,0.92) | **0.84 (0.71,1)\*** |
|  | Q3 | 0.49% | **0.80 (0.67,0.94)\*\*** | 0.87 (0.73,1.03) |
|  | Q4 | 0.46% | **0.74 (0.62,0.88) \*\*\*** | **0.83 (0.69,0.99)\*** |
|  | Q5 | 0.46% | **0.73 (0.61,0.87) \*\*\*** | **0.81 (0.67,0.98)\*** |
| Non DHA Omega 3 | IQ<sub>5</sub>R | 0.50% | 0.96(0.91,1.02) | **0.92 (0.86,0.98)\*** |
|  | Q1 | 0.55% | 1.00 | 1.00 |
|  | Q2 | 0.50% | 0.90 (0.76,1.07) | 0.89 (0.75,1.06) |
|  | Q3 | 0.47% | 0.86 (0.72,1.02) | **0.81 (0.68,0.98)\*** |
|  | Q4 | 0.49% | 0.89 (0.74,1.06) | **0.81 (0.67,0.98)\*** |
|  | Q5 | 0.49% | 0.88 (0.74,1.05) | **0.78 (0.64,0.95)\*** |

\*p<0.05, \*\*p<0.01, \*\*\*p<0.001
1. N for IQ<sub>5</sub>R models was 258,354; for individual quintile models, about 51,500 each; and for FOS use, 468,145. 2. Model 1 is adjusted for age, sex and ethnicity 3. Model 2 is adjusted for Model 1 variables plus BMI, Marital status, smoking (pack-yr), education, Townsend Deprivation index, Exercise (METs), Employment status, and Linoleic acid levels, Non-Linoleic omega-6 and Multivitamin Use. No tests of non-linearity for any of the 3 FA metrics for both outcomes were statistically significant (p>0.05 in all cases).

**Table 3.**
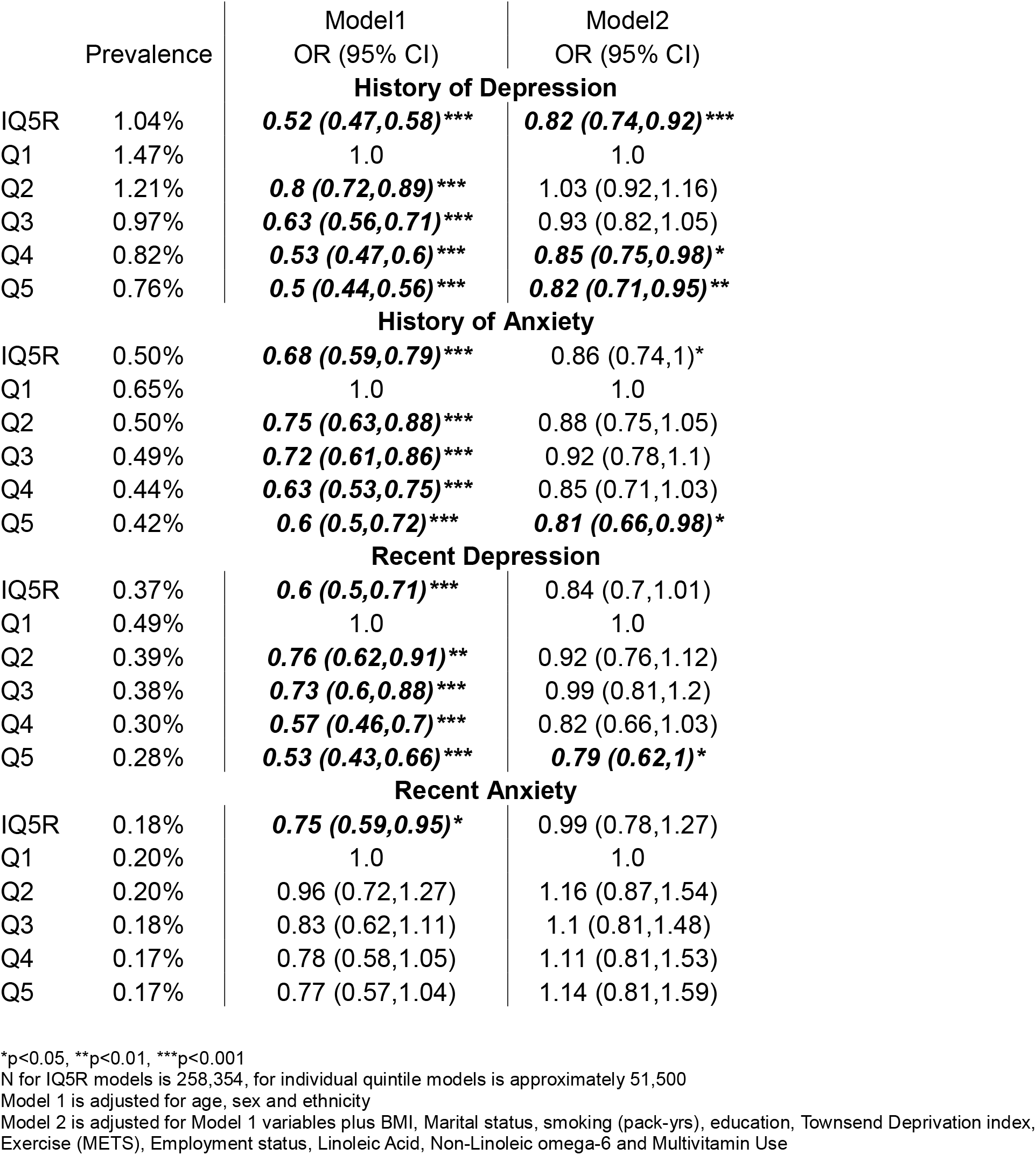
Association of the estimated Omega-3 Index with historical and recent (within the previous 12 months) depression and anxiety^1^.

For recent depression and anxiety, the associations with omega-3 measures were less clear than those seen for a history of these conditions. For recent depression, the only statistically significant findings were a 7% lower risk total omega-3 levels and a 13% lower risk for non-DHA in the linear analysis, and a 29% and 32% lower risk in Q5 vs Q1 for these two exposures, respectively. Comparing Q5 with Q1 for the eO3I there was a 21% lower risk. Plasma omega-3 levels were not significantly linked with risk for recent anxiety. Finally, no tests of non-linearity for any of the three omega-3 FA metrics by any of the outcomes were statistically significant.

### Associations between fish oil supplement use and outcomes

For FOS use, there were significant inverse relationships with risk for a history of depression and anxiety, and with risk for recent anxiety. HRs from Model 2 were 9%, 10%, and 20% lower in FOS users, respectively (**Table 4**).

**Table 4.** Association of omega-3 fatty acid levels with recent (within the previous 12 months) depression and anxiety^1^.

|  |  | Recent Depression |  |  |
| --- | --- | --- | --- | --- |
|  |  | Prevalence | Model 1<sup>2</sup><br>OR (95% CI) | Model 2<sup>3</sup><br>OR (95% CI) |
| DHA | IQ<sub>5</sub>R | 0.37% | **0.71 (0.63,0.79)\*\*\*** | 0.92 (0.82,1.04) |
|  | Q1 | 0.49% | 1.00 | 1.00 |
|  | Q2 | 0.40% | **0.79 (0.65,0.95)\*** | 1.00 (0.82,1.22) |
|  | Q3 | 0.37% | **0.72 (0.59,0.87) \*\*\*** | 1.02 (0.83,1.26) |
|  | Q4 | 0.28% | **0.53 (0.43,0.66) \*\*\*** | 0.84 (0.66,1.06) |
|  | Q5 | 0.29% | **0.55 (0.44,0.68) \*\*\*** | 0.89 (0.70,1.14) |
| Total omega 3 | IQ<sub>5</sub>R | 0.37% | **0.90 (0.86,0.94) \*\*\*** | **0.93 (0.89,0.98)\*\*** |
|  | Q1 | 0.45% | 1.00 | 1.00 |
|  | Q2 | 0.40% | 0.88(0.73,1.07) | 0.98 (0.81,1.19) |
|  | Q3 | 0.38% | 0.84(0.69,1.02) | 0.94 (0.77,1.15) |
|  | Q4 | 0.33% | **0.72 (0.58,0.88)\*\*** | 0.84 (0.68,1.04) |
|  | Q5 | 0.27% | **0.60 (0.48,0.74) \*\*\*** | **0.71 (0.56,0.9)\*\*** |
| Non DHA Omega 3 | IQ<sub>5</sub>R | 0.37% | **0.91 (0.85,0.97)\*\*** | **0.87 (0.81,0.94)\*\*\*** |
|  | Q1 | 0.42% | 1.00 | 1.00 |
|  | Q2 | 0.40% | 0.97 (0.8,1.18) | 0.96 (0.79,1.17) |
|  | Q3 | 0.38% | 0.95 (0.78,1.15) | 0.90 (0.73,1.11) |
|  | Q4 | 0.33% | **0.81 (0.66,1.00)\*** | **0.74 (0.59,0.93)\*\*** |
|  | Q5 | 0.30% | **0.75 (0.61,0.94)\*\*** | **0.68 (0.54,0.87)\*\*** |
|  |  | Recent Anxiety |  |  |
| DHA | IQ<sub>5</sub>R | 0.18% | **0.82 (0.7,0.95)\*\*** | 1.02 (0.87,1.2) |
|  | Q1 | 0.20% | 1.00 | 1.00 |
|  | Q2 | 0.18% | 0.85 (0.64,1.13) | 1.07 (0.8,1.44) |
|  | Q3 | 0.20% | 0.89 (0.67,1.18) | 1.25 (0.93,1.69) |
|  | Q4 | 0.17% | **0.73 (0.55,0.99)\*** | 1.13 (0.82,1.57) |
|  | Q5 | 0.16% | **0.71 (0.52,0.96)\*** | 1.14 (0.81,1.62) |
| Total omega 3 | IQ<sub>5</sub>R | 0.18% | 0.95 (0.89,1.01) | 0.98 (0.92,1.05) |
|  | Q1 | 0.20% | 1.00 | 1.00 |
|  | Q2 | 0.17% | 0.79 (0.59,1.06) | 0.87 (0.65,1.17) |
|  | Q3 | 0.17% | 0.80 (0.6,1.07) | 0.90 (0.67,1.21) |
|  | Q4 | 0.18% | 0.81 (0.61,1.09) | 0.96 (0.71,1.3) |
|  | Q5 | 0.19% | 0.86 (0.64,1.15) | 1.05 (0.77,1.43) |
| Non DHA Omega 3 | IQ<sub>5</sub>R | 0.18% | 0.96 (0.88,1.06) | 0.95 (0.85,1.05) |
|  | Q1 | 0.18% | 1.00 | 1.00 |
|  | Q2 | 0.19% | 1.08 (0.8,1.44) | 1.09 (0.81,1.46) |
|  | Q3 | 0.16% | 0.87 (0.64,1.19) | 0.87 (0.64,1.2) |
|  | Q4 | 0.19% | 1.03 (0.77,1.39) | 1.01 (0.74,1.39) |
|  | Q5 | 0.20% | 1.06 (0.79,1.43) | 1.05 (0.76,1.46) |

\*p<0.05, \*\*p<0.01, \*\*\*p<0.001
1. N for IQ<sub>5</sub>R models is 258,354, for individual quintile models is approximately 51,500 and for fish oil is 2. Model 1 is adjusted for age, sex and ethnicity 3. Model 2 is adjusted for Model 1 variables plus BMI, Marital status, smoking (pack-yrs), education, Townsend Deprivation index, Exercise (METs), Employment status, Linoleic Acid, Non-Linoleic omega-6 and Multivitamin Use

**Table 5.**
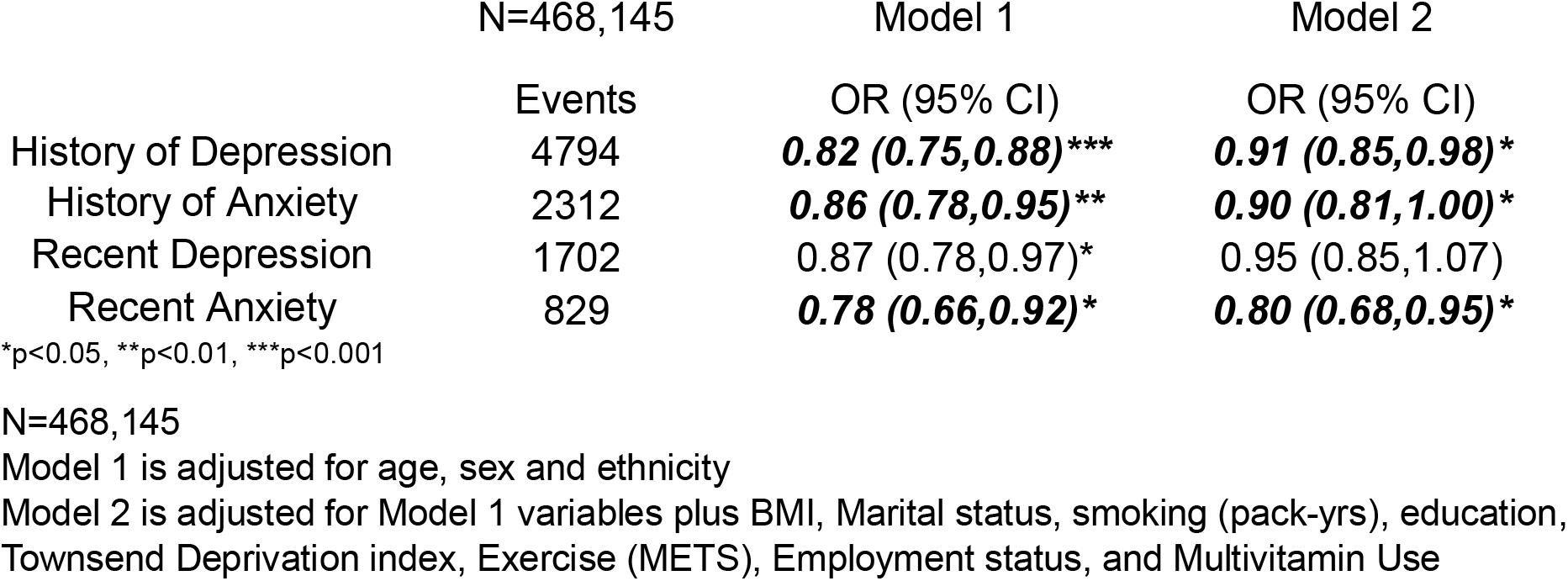
Association between the reported use of fish oil supplements (yes vs no)

|  | N=468,145 | Model 1 | Model 2 |
| --- | --- | --- | --- |
|  | Events | OR (95% CI) | OR (95% CI) |
| History of Depression | 4794 | **0.82 (0.75,0.88)\*\*\*** | **0.91 (0.85,0.98)\*** |
| History of Anxiety | 2312 | **0.86 (0.78,0.95)\*\*** | **0.90 (0.81,1.00)\*** |
| Recent Depression | 1702 | 0.87 (0.78,0.97)* | 0.95 (0.85,1.07) |
| Recent Anxiety | 829 | **0.78 (0.66,0.92)\*** | **0.80 (0.68,0.95)\*** |

\*p<0.05, \*\*p<0.01, \*\*\*p<0.001
N=468,145
Model 1 is adjusted for age, sex and ethnicity
Model 2 is adjusted for Model 1 variables plus BMI, Marital status, smoking (pack-yrs), education, Townsend Deprivation index, Exercise (METS), Employment status, and Multivitamin Use

## DISCUSSION

The purpose of this study was to examine the associations of plasma levels of omega-3 PUFAs and FOS use with depression and anxiety, both historically and concurrently. Inverse associations were generally found for the historical relationships for both of these mental disorders, but weaker or non-existent links were seen with recent disease, though in some cases risk estimates were similar to historical disease but not statistically significant. This is in part owing to the lower prevalence (hence low power) of these conditions compared with a lifetime history (about 0.37% vs 1% for depression, and 0.18% vs 0.5% for anxiety).

Other recent studies have generally supported our findings. For example, Zhang et al[11] reported that EPA intake was associated with lower risk for depressive symptoms in about 31,000 US adults based on data from the National Health And Nutrition Examination Surveys (NHANES) 2005 through 2018.

In a RCT of 71 adolescents with depression, Li et al.[12] found that adding omega-3 supplementation to Paxil treatment significantly improved depressive symptoms over 12 weeks. In older adults, a meta-analysis of five trials on depression in patients with mild cognitive impairment and/or dementia found that DHA but not EPA significantly reduced depressive symptoms in patients with dementia; however, EPA reduced depression in patients with mild cognitive impairment[13]. A study from the Cooper Center Longitudinal Study[14] found similar cross-sectional associations with depression and omega-3 PUFA levels (DHA, not EPA) and with FOS use as observed here.

As noted earlier, a meta-analysis of RCTs by Su et al.[7] found a beneficial effect of omega-3 PUFA supplementation, consistent with the observations made here. Yang et al. studied the effects of omega-3 PUFA supplementation on anxiety in first-diagnosed patients with depression being treated with venlafaxine (Effexor).[15] Although anxiety was reduced significantly compared with placebo (based on scores from the Hamilton Anxiety Scale), the reduction in scores did not correlate with changes in erythrocyte omega-3 PUFA levels[16].

### Mechanistic considerations

Omega-3 PUFAs are well-known for their ability to down regulate systemic inflammation[17]. In a mouse model of depression coupled with inflammation, both EPA and DHA provided as phospholipids alleviate depression-like behaviors. The proposed mechanisms of action related to beneficial effects of these omega-3 PUFAs on immune regulation, neuroinflammation, the hypothalamic-pituitary-adrenal axis and monoamine systems.[18]

### Strengths and Limitations

A strength of this study was the reliance on electronic medical record data instead of self-report. Although our prevalence rates were lower than they would have been had we included self-reported data, our confidence that these conditions were real (and probably more serious) was greater using physician diagnosed disorders. Obviously, the size of the UKBB cohort was a strength as was relying on biomarkers (primarily) instead of dietary intake questionnaires; reported FOS use, although an imprecise exposure metric, has been commonly used in UKBB studies[19-23] linking FOS use with disease outcomes. Weakness included the cross sectional and historical nature of the study in which reverse causation cannot be ruled out.

### Conclusions

This cross-sectional analysis of plasma omega-3 status and historical and recent depression and anxiety provides additional evidence for a favorable effect of these unique marine FAs in the etiology of these common mental disorders. Future studies should explore the prospective relationships between omega-3 PUFA biomarkers and incident depression and anxiety.

## Declaration of Generative AI in Scientific Writing

The authors declare that they made no use of generative artificial intelligence (AI) and AI-assisted technologies in the process of writing this paper.

## Authorship contributions

WSH (conception, first draft); PPD, RWG and SHM (conception, critical review); NLT (critical review and biostatistical support); JW (biostatistical support).

## Data Availability

All data produced in the present study are available upon reasonable request to the authors but the requester must also obtain permission from the UKBB for the use of these data.

---

**Supplemental Table 1.**
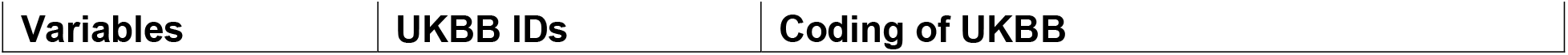
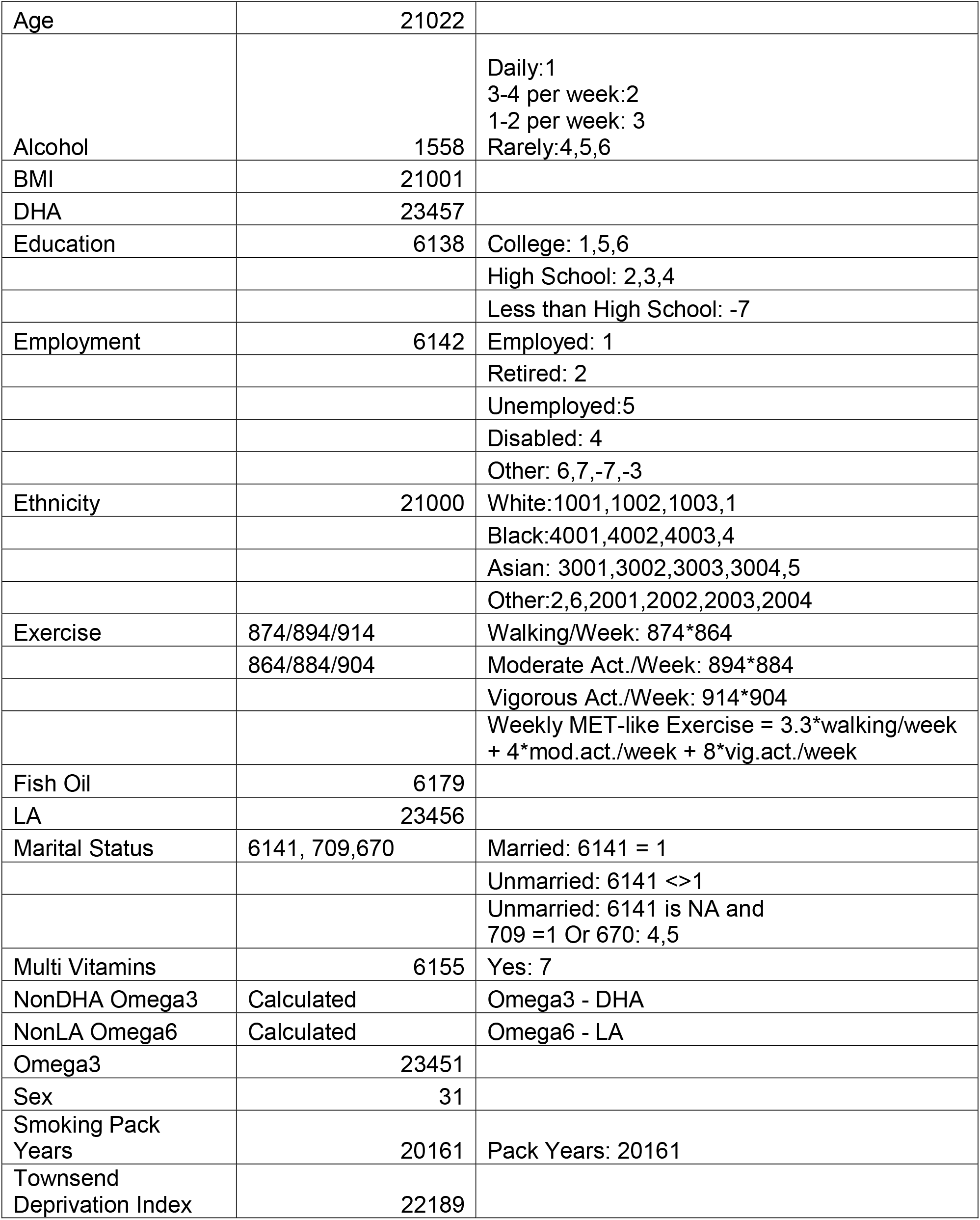

**Supplemental Table 2.**
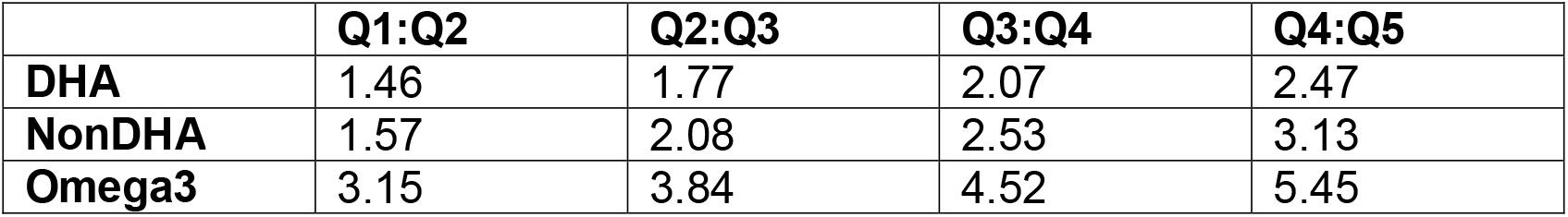
Quintile cutoffs. Values are percent of total plasma fatty acids.

