## Supplemental tables 1 and 2 for "Associations of plasma omega-3 fatty acid levels and reported fish oil supplement use with depression and anxiety: A cross-sectional analysis from the UK Biobank"

| **Supplemental Table 1** | | |
| --- | --- | --- |
| **Variables** | **UKBB IDs** | **Coding of UKBB** |
| Age | 21022 |  |
| Alcohol | 1558 | Daily:1 3-4 per week:2 1-2 per week: 3 Rarely:4,5,6 |
| BMI | 21001 |  |
| DHA | 23457 |  |
| Education | 6138 | College: 1,5,6 |
|  |  | High School: 2,3,4 |
|  |  | Less than High School: -7 |
| Employment | 6142 | Employed: 1 |
|  |  | Retired: 2 |
|  |  | Unemployed:5 |
|  |  | Disabled: 4 |
|  |  | Other: 6,7,-7,-3 |
| Ethnicity | 21000 | White:1001,1002,1003,1 |
|  |  | Black:4001,4002,4003,4 |
|  |  | Asian: 3001,3002,3003,3004,5 |
|  |  | Other:2,6,2001,2002,2003,2004 |
| Exercise | 874/894/914 | Walking/Week: 874*864 |
|  | 864/884/904 | Moderate Act./Week: 894*884 |
|  |  | Vigorous Act./Week: 914*904 |
|  |  | Weekly MET-like Exercise = 3.3*walking/week + 4*mod.act./week + 8*vig.act./week |
| Fish Oil | 6179 |  |
| LA | 23456 |  |
| Marital Status | 6141, 709,670 | Married: 6141 = 1 |
|  |  | Unmarried: 6141 <>1 |
|  |  | Unmarried: 6141 is NA and 709 =1 Or 670: 4,5 |
| Multi Vitamins | 6155 | Yes: 7 |
| NonDHA Omega3 | Calculated | Omega3 - DHA |
| NonLA Omega6 | Calculated | Omega6 - LA |
| Omega3 | 23451 |  |
| Sex | 31 |  |
| Smoking Pack Years | 20161 | Pack Years: 20161 |
| Townsend Deprivation Index | 22189 |  |

**Supplemental Table 2. Quintile cutoffs. Values are percent of total plasma fatty acids.**

|  | **Q1:Q2** | **Q2:Q3** | **Q3:Q4** | **Q4:Q5** |
| --- | --- | --- | --- | --- |
| **DHA** | 1.46 | 1.77 | 2.07 | 2.47 |
| **NonDHA** | 1.57 | 2.08 | 2.53 | 3.13 |
| **Omega3** | 3.15 | 3.84 | 4.52 | 5.45 |
